# Monocyte-chemoattractant protein-1 Levels in Human Atherosclerosis Associate with Plaque Vulnerability

**DOI:** 10.1101/2020.09.04.20187955

**Authors:** Marios K. Georgakis, Sander W. van der Laan, Yaw Asare, Joost M. Mekke, Saskia Haitjema, Arjan H. Schoneveld, Dominique P.V. de Kleijn, Gert J. de Borst, Gerard Pasterkamp, Martin Dichgans

**Author notes:** Contributed equally (Marios K. Georgakis, Sander W. van der Laan). **Corresponding author:** Martin Dichgans, MD Institute for Stroke and Dementia Research, Klinikum der Universität München Ludwig-Maximilians-University (LMU), Munich, Germany Feodor-Lynen-Str. 17, 81377 Munich, Germany.

## Abstract

Monocyte chemoattractant protein-1 (MCP-1) recruits monocytes to the atherosclerotic plaque. While experimental,^1–6^ genetic,^7^ and observational^8,9^ data support a key role of MCP-1 in atherosclerosis, the translational potential of targeting MCP-1 signaling for lowering vascular risk is limited by the lack of data on plaque MCP-1 activity in human atherosclerosis. Here, we measured MCP-1 levels in human plaque samples from 1,199 patients undergoing carotid endarterectomy and explored associations with histopathological, molecular, and clinical features of plaque vulnerability. MCP-1 plaque levels were associated with histopathological hallmarks of plaque vulnerability (large lipid core, low collagen, high macrophage burden, low smooth muscle cell burden, intraplaque hemorrhage) as well as molecular markers of plaque inflammation and matrix turnover, clinical plaque instability, and periprocedural stroke during plaque removal. Collectively, our findings highlight a role of MCP-1 in human plaque vulnerability and suggest that interfering with MCP-1 signaling in patients with established atherosclerosis could lower vascular risk.

## Introduction

Inflammatory mechanisms are critically involved in the pathogenesis of atherosclerosis.^10,11^ Recent clinical trials on cardiovascular prevention in patients with symptomatic atherosclerotic disease have demonstrated a benefit of anti-inflammatory treatment on top of standard therapy.^12–14^ These trials also highlighted the importance of targeting specific inflammatory pathways.^12,14–16^ So far, translational efforts mostly focused on the inflammasome- IL1β- IL6-axis.^17^ Yet, experimental and genetic studies place emphasis on pro-inflammatory mechanisms in atherosclerosis beyond this axis, as has specifically been shown for the chemokine system.^16,18^

Monocyte chemoattractant protein-1 (MCP-1), the prototypical CC family chemokine, attracts monocytes to sites of inflammation^19^ including the atherogenic arterial wall.^20–22^ Mice lacking MCP-1 or its receptor CCR2 are protected from atherosclerosis and pharmacological inhibition of the MCP-1/CCR2 axis reduces plaque size in experimental atherosclerosis.^1–6^ Recent genetic and observational data from humans further support associations of circulating MCP-1 levels with the risk of stroke and coronary artery disease.^7–9^ Yet, the translational potential of targeting the MCP-1/CCR2 pathway in human atherosclerosis remains elusive. Specifically, it remains unknown, whether MCP-1 activity within human plaques is causally involved in atheroprogression. To determine the potential clinical utility of targeting MCP-1, it would be critical to clarify associations between MCP-1 levels within plaques and features of plaque vulnerability and instability that underlie the occurrence of clinical events including stroke and myocardial infarction.

Here, we analyzed carotid plaque samples from > 1000 individuals, who underwent endarterectomy for treatment of asymptomatic or symptomatic carotid stenosis. Our aims were to explore associations of plaque MCP-1 levels with: (i) histopathological features of plaque vulnerability (lipid core, collagen content, macrophage burden, smooth muscle cell (SMC) burden, intraplaque hemorrhage); (ii) plaque inflammation and matrix turnover as assessed by the plaque levels of inflammatory cytokines and metalloproteinase activity; (iii) clinical plaque instability, as defined by a symptomatic plaque causing an acute cerebrovascular event; and (iv) major adverse vascular events occurring after plaque removal (**Figure 1A**).

**Figure 1.**
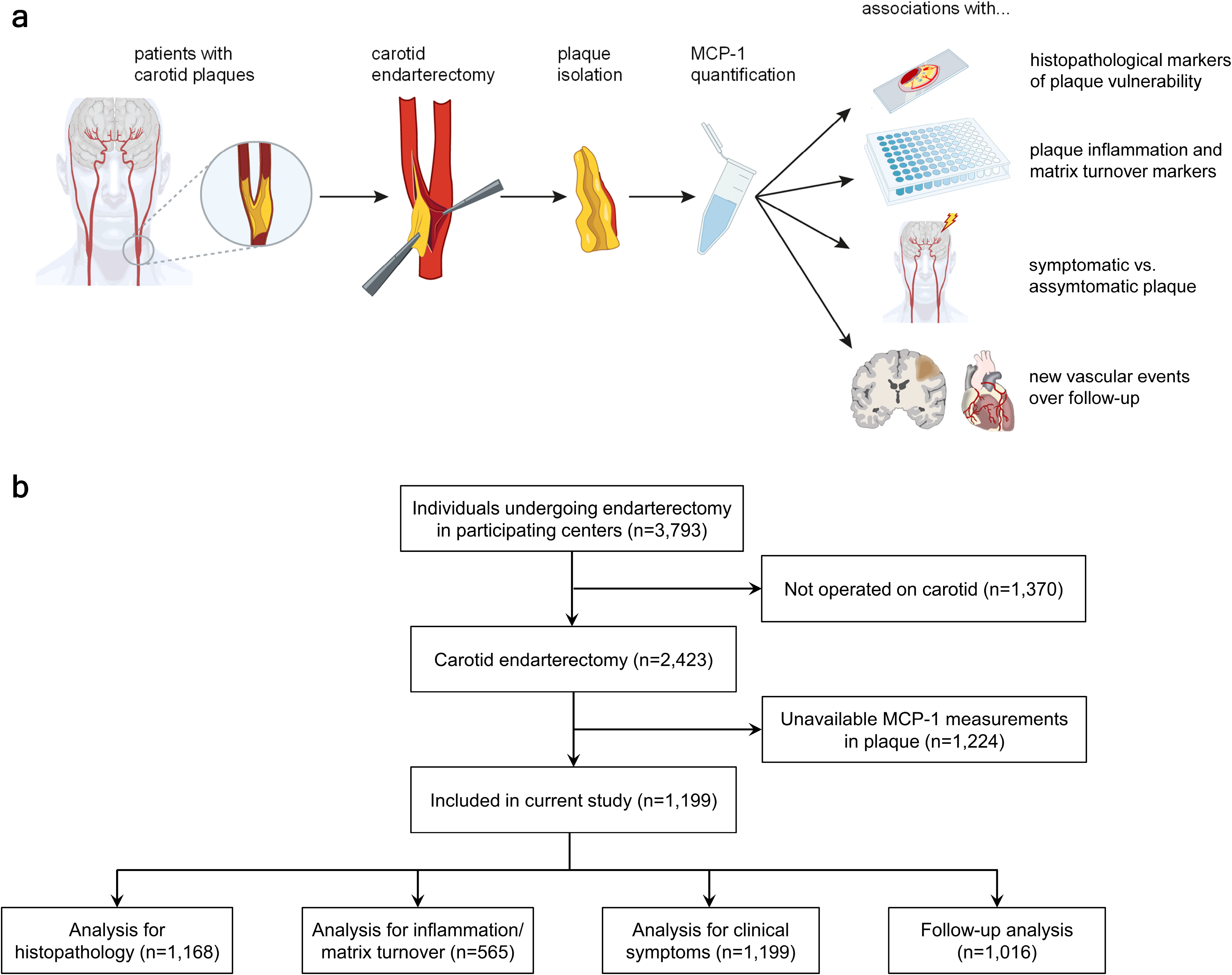
Study design. (**a**) Graphical illustration of the study design. (**b**) Flowchart detailing the number of individuals included in the respective analyses. Also shown are the number of individuals excluded from the current analyses and the reasons for exclusion.

## Results

A total of 1,199 patients from the Athero-Express Biobank, who had undergone carotid endarterectomy and had available MCP-1 levels in carotid plaques, were included in the current analysis (mean age 68.6±9.1 years, 36.3% females) (**Figure 1B, Table S1)**. MCP-1 levels in carotid plaques were higher among men compared to women, while there was no association with age or vascular risk factors including blood pressure levels, LDL-cholesterol levels, diabetes, smoking, BMI, and kidney function (**Figure S1**).

First, we explored associations of MCP-1 levels in the plaque with histopathological features of plaque vulnerability (**Figure 2**). MCP-1 levels were significantly associated with all five hallmarks of plaque vulnerability: a large lipid core (>10%), lower collagen content (no/minor), higher macrophage burden (moderate/heavy), lower SMC burden (no/minor), and presence of intraplaque hemorrhage (**Figures 2A-3B; Table S2**). Similar associations were obtained when considering macrophage and SMC burden as continuous traits (**Table S2**).

**Figure 2.**
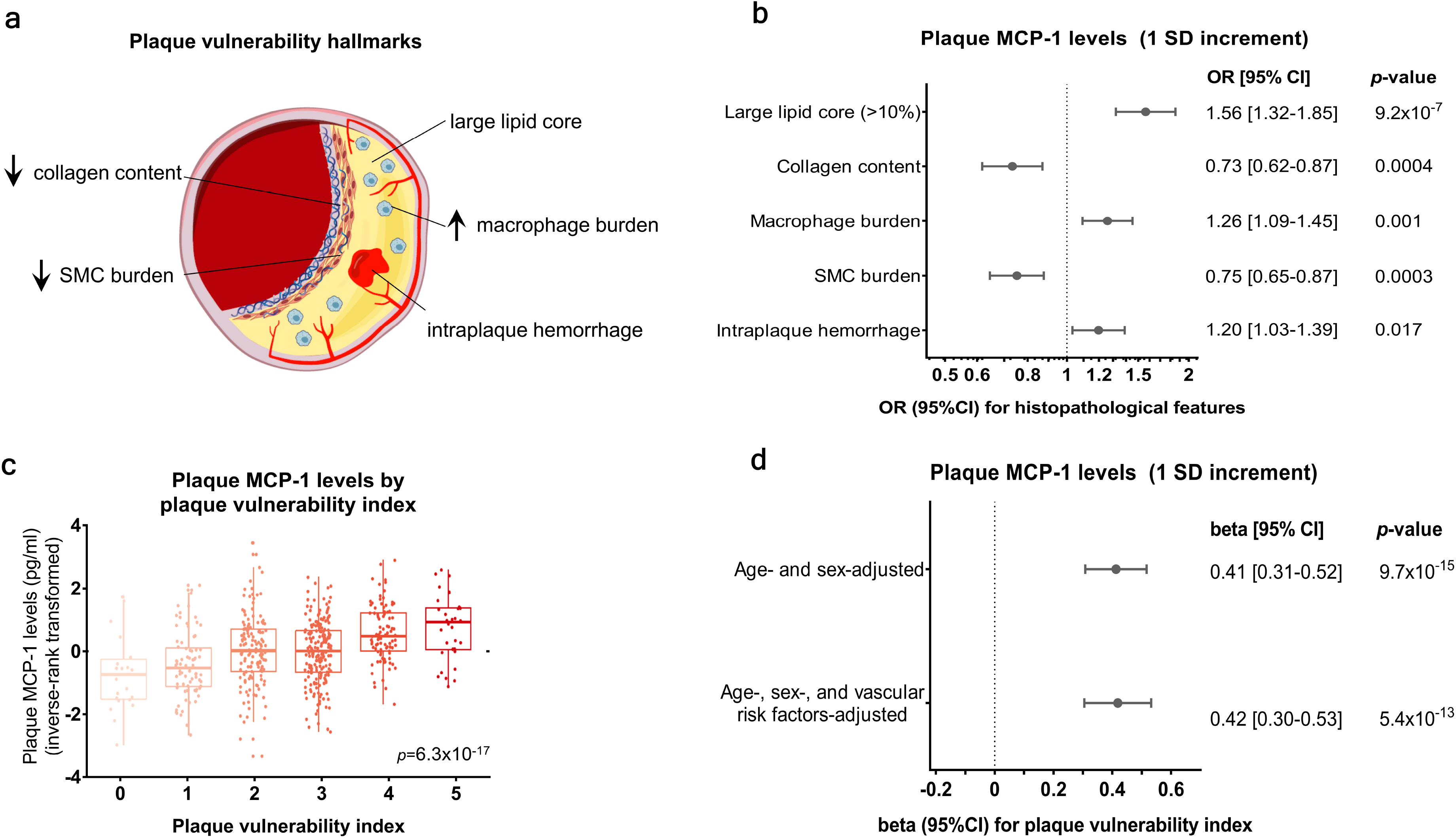
Plaque MCP-1 levels associate with histopathological hallmarks of plaque vulnerability. (**a**) Graphical depiction of histopathological features of a vulnerable plaque. (**b**) Associations of plaque MCP-1 levels (1-SD increment) with the individual vulnerability features (binary traits) as derived from logistic regression analyses (adjusted for age, sex, hypertension, diabetes, current smoking, LDL-C levels at time of operation, use of lipid-lowering agents, use of antiplatelet agents, estimated glomerular filtration rate, body mass index, history of cardiovascular disease, and grade of stenosis). Shown are Odds Ratios (OR) and error bars correspond to their 95% confidence intervals (CI). (**c**) Plaque MCP-1 levels (inverse-rank transformed) in study participants across plaque vulnerability index scores (p-value derived from Kruskal-Wallis test). Shown are the median values (central line), the upper and lower quartiles (box limits) and the 1.5x interquartile range (whiskers). (**d**) Multivariable associations between MCP-1 levels in plaque (1 SD-increment) with the composite plaque vulnerability index score, as derived from ordinal regression analyses (Model 1 adjusted for age and sex, Model 2 additionally adjusted for the abovementioned vascular risk factors). Shown are beta coefficients and the error bars correspond to their 95%CI.MCP-1 plaque levels are inverse-rank transformed in all analyses.

When combining the five hallmark features of plaque vulnerability traits in a validated aggregate vulnerability index,^23,24^ there was a dose-response relationship in that plaque MCP-1 levels were gradually higher among individuals with a higher score (ranging from 0-5, p = 6.3×10^−17^, **Figure 2C**). In models adjusting for age and sex (model 1), as well as age, sex, and vascular risk factors (model 2), plaque MCP-1 levels were strongly and independently associated with a higher vulnerability index (Model 2: beta 0.42, 95%CI: 0.30-0.53, p = 5.4×10^−13^, **Figure 2D**).

To explore the mechanisms underlying these associations we then examined whether plaque MCP-1 is associated with plaque inflammation and matrix turnover. Thus, we examined the age- and sex-adjusted associations of plaque MCP-1 levels with multiple cytokines and with metalloproteinase activity in carotid plaques (**Figure 3**). We found significant associations between MCP-1 plaque levels and several cytokines involved in inflammatory cell recruitment. Specifically, we found associations with higher levels of the chemokines IL-8, PARC, TARC, and RANTES, as well with ICAM-1, an adhesion molecule involved in transendothelial leukocyte migration^25,26^ (all FDR-adjusted p-value < 0.05 to account for multiple comparisons). Higher MCP-1 levels were further associated with higher levels of VEGF-A, a key driver of plaque neovascularization,^27^ and with higher activity of the matrix metalloproteinases MMP-8, and MMP-9, thus supporting a role of MCP-1 in matrix turnover.^28^ Similar results were obtained when further adjusting for vascular risk factors, although associations with PARC did not remain statistically significant (Model 2, **Table S3**).

**Figure 3.**
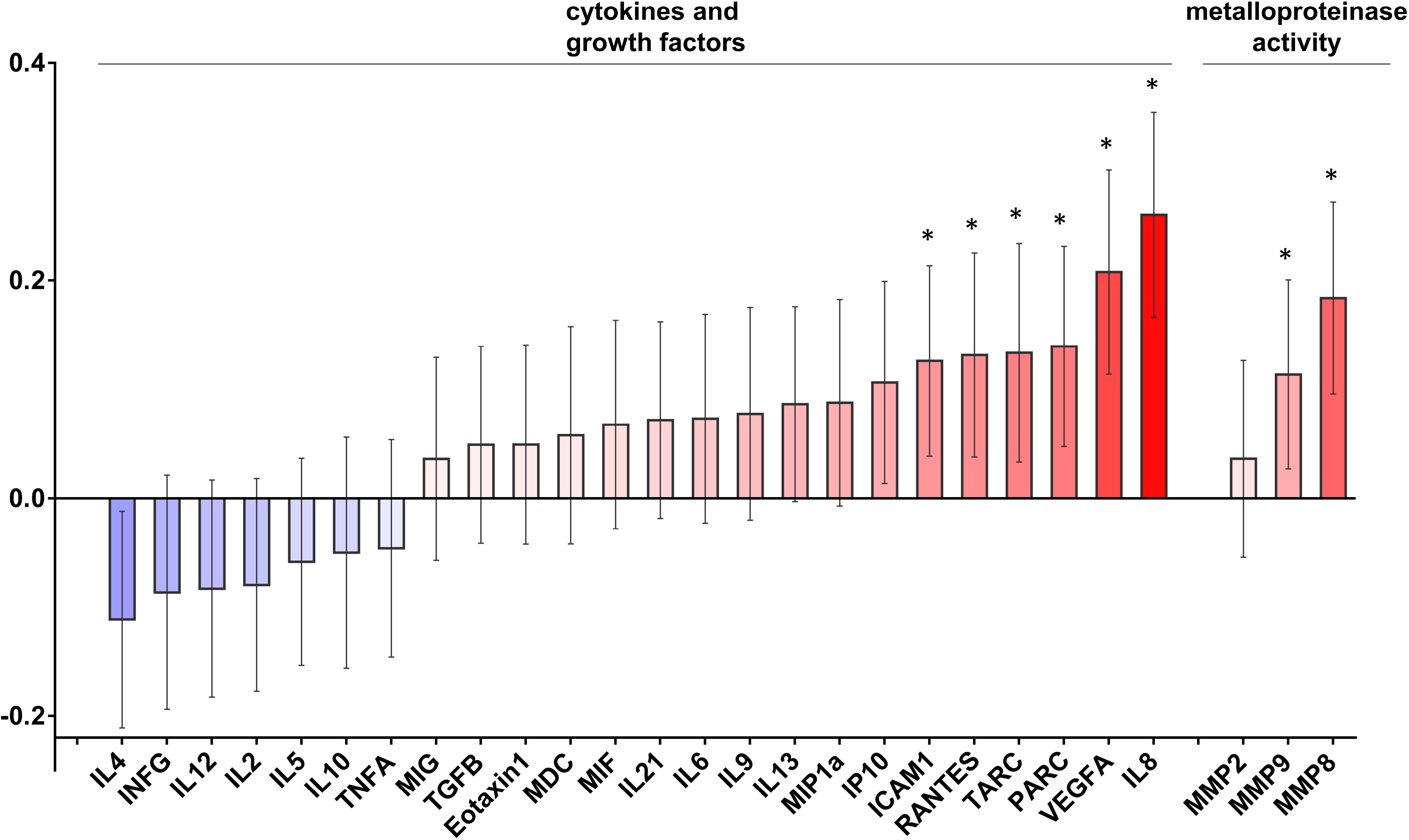
Plaque MCP-1 levels associate with markers of plaque inflammation and matrix turnover. Shown are the beta coefficients and 95% confidence intervals (95%CI) derived from linear regression models for 1-SD increment in plaque MCP-1 levels (inverse- rank transformed) adjusted for age and sex. Stars indicate statistically significant results (false discovery rate-adjusted p-value < 0.05).

We further explored associations between plaque MCP-1 levels and clinical plaque instability. MCP-1 levels in the plaque were higher among individuals with a symptomatic plaque (that had caused an acute cerebrovascular event) compared to individuals with asymptomatic plaques (p = 0.0001, **Figure 4A**). Following adjustments for age and sex, one SD increment in MCP-1 levels in the plaque was associated with higher odds for a symptomatic vs. asymptomatic plaque (OR: 1.31, 95%CI: 1.07-1.60, p = 0.008). These associations remained significant in a model additionally adjusting for vascular risk factors (OR: 1.36, 95%CI: 1.09-1.69, p = 0.006, **Figure 4B**).

**Figure 4.**
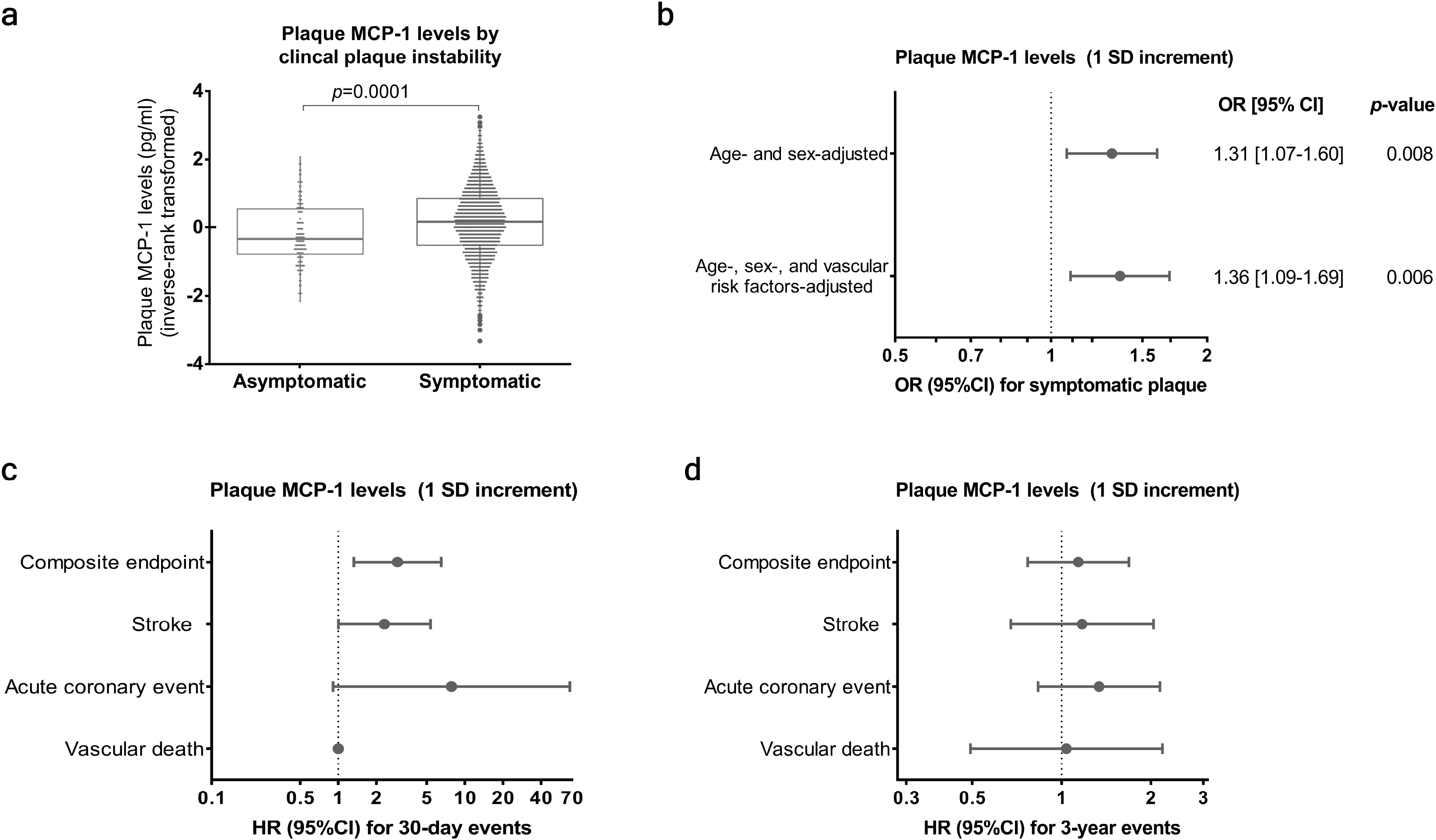
Plaque MCP-1 levels associate with symptomatic versus asymptomatic plaque and periprocedural events after plaque removal. (**a**) Plaque MCP-1 levels (inverse-rank transformed) in patients with symptomatic versus asymptomatic plaques (pvalue derived from Mann-Whitney U-test). Shown are the median values (central line), the upper and lower quartiles (box limits) and the 1.5x interquartile range (whiskers) (**b**) Multivariable associations between MCP-1 plaque levels (1 SD-increment) with symptomatic plaque as derived from logistic regression analyses (vascular risk factors in the second model include hypertension, diabetes, current smoking, LDL-C levels at time of operation, use of lipid-lowering agents, use of antiplatelet agents, estimated glomerular filtration rate, body mass index, history of cardiovascular disease, and grade of stenosis). Shown are Odds Ratios (OR) and error bars correspond to their 95% confidence intervals (CI). (**c, d**) Associations of plaque MCP-1 levels (1-SD increment) with new adverse vascular events within (**c**) 30 days (periprocedural events) and (**d**) 3 years after surgery, as derived from Cox regression analyses (adjusted for age, sex, and the abovementioned vascular risk factors). Shown are Hazard Ratios (OR) derived from Cox regression analyses and error bars correspond to their 95%CI. MCP-1 plaque levels are inverse-rank transformed in all analyses.

As a last step, we explored associations between MCP-1 levels in the plaque with vascular events after carotid endarterectomy. Specifically, we looked at events occurring within the first 30 days after surgery mostly reflective of periprocedural complications, and at events occurring up to 3 years after surgery (mean follow-up 2.3 years). Plaque MCP-1 levels (1-SD increment) were associated with a higher risk of major adverse vascular events within 30-days after surgery (HR: 2.94, 95%CI: 1.32-6.50, p = 0.007, **Figure 4C**) independently of age, sex, and vascular risk factors (**Table S4**) whereas there was no such association across the entire 3-year period (HR: 1.14, 95%CI: 0.77-1.68, p = 0.519, **Figure 4D, Table S4**). MCP-1 plaque levels were further associated with a higher 30-day risk of stroke (HR: 2.32, 95%CI: 1.00-5.36, p = 0.049, **Figure 4C**).

## Discussion

The present study of 1,199 patients undergoing carotid endarterectomy demonstrated strong associations between plaque MCP-1 levels with multiple features of plaque vulnerability. We found associations between plaque MCP-1 levels and histopathological hallmarks of plaque vulnerability (larger lipid core, low collagen content, high macrophage burden, low SMC burden, intraplaque hemorrhage). We further found higher plaque MCP-1 levels to be associated with higher levels of inflammatory cytokines and higher activity of metalloproteinases within plaques indicating associations with a pro-inflammatory plaque profile and matrix turnover. MCP-1 levels within plaques were higher among individuals with symptomatic plaques, as compared to asymptomatic plaques. Moreover, plaque MCP-1 levels were associated with adverse peri-procedural vascular events (time frame: 30 days after plaque removal) independently of traditional vascular risk factors. Collectively, these findings emphasize the importance of plaque MCP-1 levels in plaque vulnerability in human atherosclerosis.

While the role of MCP-1 in early stages of atherogenesis through monocyte recruitment in the plaque has been well-established,^20–22^ its role in more advanced stages of atherosclerosis remained unknown. The associations reported here between MCP-1 levels in the plaque with all five hallmarks of vulnerable plaques, as well as with a composite vulnerability index, suggest that MCP-1 is involved in mechanisms related to plaque instability in patients. Our finding of significant associations between plaque MCP-1 levels and symptomatic plaques further support this notion. Importantly, these associations were independent of conventional vascular risk factors, thus suggesting that MCP-1 signaling might contribute to plaque instability on top of established targets for secondary prevention such as LDL-cholesterol, blood pressure, and diabetes.

Our current findings on MCP-1 levels in carotid endarterectomy samples complement our recent work showing that circulating MCP-1 levels associate with ischemic stroke, coronary artery disease, and vascular death.^7–9^ Whether MCP-1 levels within atherosclerotic plaques are reflective of MCP-1 activity and active monocyte recruitment remains unknown. Yet, our finding of an association between MCP-1 levels in the plaque with multiple other cell-recruiting chemokines with a proven role in atherosclerosis,^29–32^ as well as with macrophage burden in the plaque strongly suggest an association with inflammatory cell recruitment

While MCP-1 levels within plaques were not associated with future vascular events up to 3 years after plaque removal, we found significant associations with periprocedural events, mainly stroke, occurring within the first 30 days after the procedure. Periprocedural strokes during carotid endarterectomy might have multiple causes, but have been shown to frequently originate from plaque thrombosis and embolization.^33^ Although the mechanisms underlying the observed associations remain unknown, they might relate to the higher risk of periprocedural microembolization previously reported to be more common in patients with features of vulnerable plaques.^34,35^

To our knowledge, there has been only one small phase II trial targeting MCP-1 signaling in the context of human atherosclerosis. Among 108 patients with vascular risk factors and high CRP levels, treatment with a single intravenous infusion of a humanized monoclonal antibody against the receptor of MCP-1 (CCR2), led to significant reductions in CRP levels.^36^ Of note, the residual risk for clinical events among individuals with carotid plaques currently managed by best medical treatment (statins and anti-platelet agents) remains non-negligible.^37–39^ Our current data in conjunction with recent genetic,^7^ experimental,^1–6^ and observational^8,9^ data on MCP-1 support moving towards clinical trials that target MCP-1 signaling in populations with established atherosclerotic disease.

Our study has limitations. First, the cross-sectional nature of most analyses precludes causal inferences. For example, the association between plaque MCP-1 levels and symptomatic plaques could relate to a secondary increase in plaque MCP-1 levels following the acute clinical event.^40^ Second, plaque MCP-1 levels were available only in around half of the Athero-Express population, thus potentially leading to selection bias. Yet, the demographic characteristics of our study sample matched those of the entire Athero-Express sample (**Table S1**). Similarly, there might be selection bias due to the underrepresentation of patients with asymptomatic plaques, as they are less likely to undergo carotid endarterectomy. Third, our longitudinal analyses are limited by power due to the low number of new events in the current cohort. Fourth, the pathology and mechanisms of plaque vulnerability might differ between vascular beds and these results from carotid plaques might not be extrapolated to other atherosclerotic lesions.

In conclusion, our study shows that among individuals undergoing carotid endarterectomy, plaque MCP-1 levels are associated with histopathological hallmarks of plaque vulnerability, a pro-inflammatory plaque profile, plaque matrix turnover, clinical plaque instability, and higher risk of periprocedural events. As such, our findings provide evidence for an involvement of MCP-1 in carotid plaque instability and further complement previous evidence from epidemiological, genetic, and experimental studies supporting the translational perspective of targeting the MCP-1/CCR2 axis in atherosclerosis.

## METHODS

### Study population

We used data from the Athero-Express Biobank (http://www.atheroexpress.nl), an ongoing prospective study of patients undergoing endarterectomy for manifestations of atherosclerosis.^41^ Carotid endarterectomy was performed following recommendations by the ACAS^42^ and NASCET^43^ studies. Patients were recruited from the St. Antonius Hospital Nieuwegein and University Medical Center Utrecht in Utrecht, Netherlands between 2002 and 2019. Individuals who agreed to participate completed questionnaires about medical history and medication use prior to the operation and provided blood samples for biochemical and hematological analyses. Their plaque samples were post-operatively collected and analyzed as described below. Individuals were included in the current study on the basis of having undergone carotid endarterectomy and having available measurements of MCP-1 levels in plaque (**Figure 1**). The study protocol conforms to the Declaration of Helsinki and was approved by the ethics committee on research on humans of the University Medical Center Utrecht. All participants provided written informed consent.

### Histopathological analysis of atherosclerotic plaque composition

Following carotid endarterectomy, plaque samples were immediately transferred to the laboratory. Plaques were divided in parallel segments of 5-mm thickness perpendicular to the arterial axis and the segment with the greatest plaque burden was subjected to histopathological examination, as previously described.^34,44,45^ All stained sections were examined microscopically and digitally stored. For the purposes of the current study, we explored five plaque traits that are established hallmarks of plaque vulnerability: lipid content, collagen deposition, macrophages, smooth muscle cells, and intraplaque hemorrhage.^46,47^ Two independent observers manually scored stainings for these traits using previously defined semi-quantitative methods.^34,41,44,45^ In brief, plaque lipid content was quantified visually as a percentage of fat deposition to total plaque area with the use of H&E and picrosirius red stains; a large lipid core was defined as lipid content of > 10% of the total plaque area. Collagen deposition (picrosirius red) was manually classified as absent, minor, moderate or heavy staining along the entire luminal border. The burden of macrophages and SMCs was assessed by staining with antibodies against CD68 and α-actin, respectively, and was also manually classified into absent, minor, moderate or heavy staining. In alternative semi-automated computerized analyses, numbers of macrophages and SMCs were quantified on a continuous scale. Specifically, the stainings were scored as percentage of stained area to total plaque area (AnalySiS version 3.2, Soft Imaging GmbH, Munster, Germany).^34,44,45^ Intraplaque hemorrhage (H&E and fibrin staining) was defined as the composite of plaque bleeding at the luminal side of the plaque as a result of plaque disruption, and was classified as absent or present.

To assess the overall vulnerability features of the atherosclerotic plaque, a vulnerability index was created ranging from 0 to 5, as previously described.^23,24^ Specifically, one point was given to each plaque for the following histopathological features: a lipid core > 10%, low collagen load (no/minor), high macrophage burden (moderate/heavy), low SMC burden (no/minor), and presence of intraplaque hemorrhage.

### Quantification of plaque levels of MCP-1 and other cytokines

Segments adjacent to those used for histopathological analysis were used for protein isolation. In brief, plaques were manually grinded at −196°C and dissolved in Tris buffer according to an in-house protocol.^41^ MCP-1 concentrations (pg/mL) were quantified as part of a multiplex assay using the Luminex® platform (Austin, TX, USA) according to the manufacturer’s protocol and diagnostic laboratories’ standards at the clinical laboratory of the Wilhelmina Children’s Hospital (WKZ, Utrecht, the Netherlands).

We further quantified the following cytokines and growth factors using established platforms: IL2, IL4, IL5, IL6, IL8, IL9, IL10, IL12, TNF-α, and IFN-γ were measured in multiplex using the human FlowCytomix system from eBioscience (cat.nr.: BMS810FF) in pg/mL. IL13, IL21, MIF, MIP1a, RANTES, MIG, IP10, Eotaxin1, TARC, PARC, MDC, sICAM1, VEGFA, and TGFB were measured in simplex assays using FlowCytomix according to the manufacturer’s protocol and diagnostic laboratories’ standards at the clinical laboratory of the WKZ. Metalloproteinase (MMP) activity (MMP-2, MMP-8, MMP-9) was assessed with specific Biotrak activity assays (MMP-2 RPN-2631, MMP-8 RPN-2635, and MMP-9 RPN-2634; GE Healthcare LifeSciences, Buckinghamshire, UK). Matrix metalloproteinase levels were corrected for the total protein amount and were expressed as arbitrary units (AU). Given the different platforms used, all protein measurements were inverse-rank transformed to approach normal distributions and to ensure homogeneity in units (per 1-standard deviation [SD]).

### Symptomatic vs. asymptomatic plaque

Patients were classified as either having an asymptomatic or symptomatic carotid plaque prior to surgery, based on their answers to a structured questionnaire regarding prior medical history and a detailed review of their medical records. Patients were considered to be symptomatic if they had suffered an acute cerebrovascular event ipsilateral to the plaque within the last 6 months. Cerebrovascular events included a supratentorial ischemic stroke, a transient ischemic attack that could be attributed to ischemia in the distribution of the respective artery, an amaurosis fugax, or a central retinal artery occlusion.

### Follow-up analysis for major adverse vascular events

Patients were followed up to 3 years after surgery for potential new vascular events. The composite endpoint of any major adverse vascular event included non-fatal stroke (ischemic or hemorrhagic), non-fatal myocardial infarction, ruptured aortic aneurysm, and vascular death, defined as death of presumed vascular origin (fatal stroke, fatal myocardial infarction, sudden death, fatal aortic aneurysm rupture, fatal heart failure, other vascular death). Additional endpoints included stroke (fatal or non-fatal), acute coronary events (fatal or nonfatal myocardial infarction, unstable angina, coronary bypass or percutaneous coronary intervention, and sudden cardiac death), as well as vascular death. Outcomes occurring within the first 30 days after surgery were considered periprocedural events.^34,48,49^ All participants underwent clinical follow-up, as detailed elsewhere.^34^ Clinical endpoints were independently assessed by two clinicians at 1, 2, and 3 years after surgery through patient questionnaires, review of medical records and contact with general practitioners.

### Statistical analysis

Univariable associations between inverse-rank transformed levels of MCP-1 in the plaque with other group variables were explored using t-test or ANOVA, as appropriate. Multivariable models were used to explore associations of plaque levels of MCP-1 (1-SD increment) with (i) plaque protein levels of a panel of 24 cytokines and growth factors (thereafter called cytokines for simplicity) and activity of three metalloproteinases, (ii) histopathological plaque vulnerability phenotypes, (iii) presence of a symptomatic vs. asymptomatic plaque, and (iv) incident major adverse vascular events. Specifically, we performed multivariable logistic regression analyses for symptomatic vs. asymptomatic plaque and for binary histopathological plaque vulnerability traits, as well as linear regression analyses for plaque cytokines and for continuous histopathological plaque vulnerability traits. For the composite vulnerability index (range 0-5), we used ordinal regression analyses. For the prospective analyses for time to new major adverse vascular events, we applied Cox proportional hazard models. Model 1 adjusted for age and sex, whereas Model 2 additionally adjusted for hypertension (self-reported history or antihypertensive medication use), diabetes (defined as self-reported history or glucose-lowering medication use), smoking status (never, former, current smoker), LDL-C levels at time of operation, use of statins or other lipid-lowering drugs, use of antiplatelet agents, estimated glomerular filtration rate (eGFR),^50^ body mass index (BMI), history of cardiovascular disease (coronary artery disease, stroke, peripheral artery disease), and grade of stenosis (according to NASCET: < 70%, 70-90%, 90-99%, complete occlusion). All analyses were corrected for multiple comparisons using the false discovery rate (FDR) approach. Statistical significance threshold was set at a two-sided FDR-adjusted p-value< 0.05 across all analyses. Analyses were performed using R (v3.6.3; The R Foundation for Statistical Computing).

### Data availability

The datasets from Ahero-Express analyzed during the current study are available upon request to the corresponding author and application to Athero-Express through a Template Material/Data Transfer Agreement due to consent restriction. The code used for analyzing the data for the current study is available through GitHub under the following link: https://github.com/swvanderlaan/2020_georgakis_vanderlaan_MCP1.

## Data Availability

The datasets from Ahero-Express analyzed during the current study are available upon request to the corresponding author and application to Athero-Express through a Template Material/Data Transfer Agreement due to consent restriction. The code used for the current study is available through GitHub under the following link: https://github.com/swvanderlaan/2020_georgakis_vanderlaan_MCP1

## Acknowledgements

This project has received funding from the European Union’s Horizon 2020 research and innovation programme (666881), SVDs@target (to MD; 667375), CoSTREAM (to MD); the DFG as part of the Munich Cluster for Systems Neurology (EXC 2145 SyNergy – ID 390857198) and the CRC 1123 (B3; to MD); the Corona Foundation (to MD); the Fondation Leducq (Transatlantic Network of Excellence on the Pathogenesis of Small Vessel Disease of the Brain; to MD); the e:Med program (e:AtheroSysMed; to MD) and the FP7/2007-2103 European Union project CVgenes@target (grant agreement number Health-F2-2013-601456; to MD).

## Author contributions

MKG conceived and designed the current study, designed the statistical analysis, and wrote the first draft of the manuscript. SWvdL designed the current study, performed the statistical analysis, and revised the manuscript for intellectual content. YA designed the current study and revised the manuscript for intellectual content. JMM and SH recruited patients in Athero-Express and revised the manuscript for intellectual content. AS performed the protein measurements in Athero-Express and revised the manuscript for intellectual content. DPVdK performed the measurements of protein levels and metalloproteinase activity in Athero-Express and revised the manuscript for intellectual content. GJdB designed Athero-Express, recruited patients for the study and revised the manuscript for intellectual content. GP designed Athero-Express and revised the manuscript for intellectual content. MD designed the current study and wrote the first draft of the manuscript.

## References

1 Liehn, E.A., et al A new monocyte chemotactic protein-1/chemokine CC motif ligand-2 competitor limiting neointima formation and myocardial ischemia/reperfusion injury in mice. J Am Coll Cardiol 56, 1847–1857 (2010).

2 Bot, I., et al A novel CCR2 antagonist inhibits atherogenesis in apoE deficient mice by achieving high receptor occupancy. Sci Rep 7, 52 (2017).

3 Gu, L., et al Absence of monocyte chemoattractant protein-1 reduces atherosclerosis in low density lipoprotein receptor-deficient mice. Mol Cell 2, 275–281 (1998).

4 Boring, L., Gosling, J., Cleary, M. & Charo, I.F. Decreased lesion formation in CCR2-/- mice reveals a role for chemokines in the initiation of atherosclerosis. Nature 394, 894–897 (1998).

5 Combadiere, C., et al Combined inhibition of CCL2, CX3CR1, and CCR5 abrogates Ly6C(hi) and Ly6C(lo) monocytosis and almost abolishes atherosclerosis in hypercholesterolemic mice. Circulation 117, 1649–1657 (2008).

6 Winter, C., et al Chrono-pharmacological Targeting of the CCL2-CCR2 Axis Ameliorates Atherosclerosis. Cell Metab 28, 175–182 e175 (2018).

7 Georgakis, M.K., et al Genetically Determined Levels of Circulating Cytokines and Risk of Stroke. Circulation 139, 256–268 (2019).

8 Georgakis, M.K., et al Circulating Monocyte Chemoattractant Protein-1 and Risk of Stroke: Meta-Analysis of Population-Based Studies Involving 17 180 Individuals. Circ Res 125, 773–782 (2019).

9 Georgakis, M.K., et al Circulating monocyte chemoattractant protein-1 levels are associated with 1 cardiovascular mortality: a meta-analysis of population-based studies. JAMA Cardiology (Accepted) (2020).

10 Esenwa, C.C. & Elkind, M.S. Inflammatory risk factors, biomarkers and associated therapy in ischaemic stroke. Nat Rev Neurol 12, 594–604 (2016).

11 Libby, P., Ridker, P.M., Hansson, G.K. & Leducq Transatlantic Network on Atherothrombosis. Inflammation in atherosclerosis: from pathophysiology to practice. J Am Coll Cardiol 54, 2129–2138 (2009).

12 Ridker, P.M., et al Antiinflammatory Therapy with Canakinumab for Atherosclerotic Disease. N Engl J Med 377, 1119–1131 (2017).

13 Tardif, J.C., et al Efficacy and Safety of Low-Dose Colchicine after Myocardial Infarction. N Engl J Med 381, 2497–2505 (2019).

14 Ridker, P.M., et al Low-Dose Methotrexate for the Prevention of Atherosclerotic Events. N Engl J Med 380, 752–762 (2019).

15 Ridker, P.M. Anticytokine Agents: Targeting Interleukin Signaling Pathways for the Treatment of Atherothrombosis. Circ Res 124, 437–450 (2019).

16 Aday, A.W. & Ridker, P.M. Targeting Residual Inflammatory Risk: A Shifting Paradigm for Atherosclerotic Disease. Front Cardiovasc Med 6, 16 (2019).

17 Ridker, P.M. From C-Reactive Protein to Interleukin-6 to Interleukin-1: Moving Upstream To Identify Novel Targets for Atheroprotection. Circ Res 118, 145–156 (2016).

18 Koenen, R.R. & Weber, C. Therapeutic targeting of chemokine interactions in atherosclerosis. Nat Rev Drug Discov 9, 141–153 (2010).

19 Deshmane, S.L., Kremlev, S., Amini, S. & Sawaya, B.E. Monocyte chemoattractant protein-1 (MCP-1): an overview. J Interferon Cytokine Res 29, 313–326 (2009).

20 Lin, J., Kakkar, V. & Lu, X. Impact of MCP-1 in atherosclerosis. Curr Pharm Des 20, 4580–4588 (2014).

21 Nelken, N.A., Coughlin, S.R., Gordon, D. & Wilcox, J.N. Monocyte chemoattractant protein-1 in human atheromatous plaques. J Clin Invest 88, 1121–1127 (1991).

22 Lutgens, E., et al Gene profiling in atherosclerosis reveals a key role for small inducible cytokines: validation using a novel monocyte chemoattractant protein monoclonal antibody. Circulation 111, 3443–3452 (2005).

23 Timmerman, N., et al Family history and polygenic risk of cardiovascular disease: Independent factors associated with secondary cardiovascular events in patients undergoing carotid endarterectomy. Atherosclerosis (2020).

24 Verhoeven, B., et al Carotid atherosclerotic plaques in patients with transient ischemic attacks and stroke have unstable characteristics compared with plaques in asymptomatic and amaurosis fugax patients. J Vasc Surg 42, 1075–1081 (2005).

25 Kitagawa, K., et al Involvement of ICAM-1 in the progression of atherosclerosis in APOE-knockout mice. atherosclerosis 160, 305–310 (2002).

26 Nakashima, Y., Raines, E.W., Plump, A.S., Breslow, J.L. & Ross, R. Upregulation of VCAM-1 and ICAM-1 at atherosclerosis-prone sites on the endothelium in the ApoE-deficient mouse. Arterioscler Thromb Vasc Biol 18, 842–851 (1998).

27 Camare, C., Pucelle, M., Negre-Salvayre, A. & Salvayre, R. Angiogenesis in the atherosclerotic plaque. Redox Biol 12, 18–34 (2017).

28 Peeters, W., et al Collagenase matrix metalloproteinase-8 expressed in atherosclerotic carotid plaques is associated with systemic cardiovascular outcome. Eur Heart J 32, 2314–2325 (2011).

29 Simonini, A., et al IL-8 is an angiogenic factor in human coronary atherectomy tissue. Circulation 101, 1519–1526 (2000).

30 Veillard, N.R., et al Antagonism of RANTES receptors reduces atherosclerotic plaque formation in mice. Circ Res 94, 253–261 (2004).

31 Weber, C., et al CCL17-expressing dendritic cells drive atherosclerosis by restraining regulatory T cell homeostasis in mice. J Clin Invest 121, 2898–2910 (2011).

32 Orekhov, A.N., et al Tumor Necrosis Factor-alpha and C-C Motif Chemokine Ligand 18 Associate with Atherosclerotic Lipid Accumulation In situ and In vitro. Curr Pharm Des 24, 2883–2889 (2018).

33 Jacobowitz, G.R., et al Causes of perioperative stroke after carotid endarterectomy: special considerations in symptomatic patients. Ann Vasc Surg 15, 19–24 (2001).

34 Hellings, W.E., et al Composition of carotid atherosclerotic plaque is associated with cardiovascular outcome: a prognostic study. Circulation 121, 1941–1950 (2010).

35 Verhoeven, B.A., et al Carotid atherosclerotic plaque characteristics are associated with microembolization during carotid endarterectomy and procedural outcome. Stroke 36, 1735–1740 (2005).

36 Gilbert, J., et al Effect of CC chemokine receptor 2 CCR2 blockade on serum C-reactive protein in individuals at atherosclerotic risk and with a single nucleotide polymorphism of the monocyte chemoattractant protein-1 promoter region. Am J Cardiol 107, 906–911 (2011).

37 Spence, J.D., et al Effects of intensive medical therapy on microemboli and cardiovascular risk in asymptomatic carotid stenosis. Arch Neurol 67, 180–186 (2010).

38 Inzitari, D., et al The causes and risk of stroke in patients with asymptomatic internal-carotid-artery stenosis. North American Symptomatic Carotid Endarterectomy Trial Collaborators. N Engl J Med 342, 1693–1700 (2000).

39 Nadareishvili, Z.G., Rothwell, P.M., Beletsky, V., Pagniello, A. & Norris, J.W. Long-term risk of stroke and other vascular events in patients with asymptomatic carotid artery stenosis. Arch Neurol 59, 1162–1166 (2002).

40 Roth, S., et al Brain-released alarmins and stress response synergize in accelerating atherosclerosis progression after stroke. Sci Transl Med 10 (2018).

41 Verhoeven, B.A., et al Athero-express: differential atherosclerotic plaque expression of mRNA and protein in relation to cardiovascular events and patient characteristics. Rationale and design. Eur J Epidemiol 19, 1127–1133 (2004).

42 Toole, J.F. ACAS recommendations for carotid endarterectomy. ACAS Executive Committee. Lancet 347, 121 (1996).

43 Coyne, T.J. & Wallace, M.C. Surgical referral for carotid artery stenosis--the influence of NASCET. North American Symptomatic Carotid Endarterectomy Trial. Can J Neurol Sci 21, 129–132 (1994).

44 Hellings, W.E., et al Intraobserver and interobserver variability and spatial differences in histologic examination of carotid endarterectomy specimens. J Vasc Surg 46, 1147–1154 (2007).

45 Meeuwsen, J.A.L., et al Circulating CD14(+)CD16(−) classical monocytes do not associate with a vulnerable plaque phenotype, and do not predict secondary events in severe atherosclerotic patients. J Mol Cell Cardiol 127, 260–269 (2019).

46 Finn, A.V., Nakano, M., Narula, J., Kolodgie, F.D. & Virmani, R. Concept of vulnerable/unstable plaque. Arterioscler Thromb Vasc Biol 30, 1282–1292 (2010).

47 Bentzon, J.F., Otsuka, F., Virmani, R. & Falk, E. Mechanisms of plaque formation and rupture. Circ Res 114, 1852–1866 (2014).

48 Randomised trial of endarterectomy for recently symptomatic carotid stenosis: final results of the MRC European Carotid Surgery Trial (ECST). Lancet 351, 1379–1387 (1998).

49 Barnett, H.J., et al Benefit of carotid endarterectomy in patients with symptomatic moderate or severe stenosis. North American Symptomatic Carotid Endarterectomy Trial Collaborators. N Engl J Med 339, 1415–1425 (1998).

50 Levey, A.S., et al A new equation to estimate glomerular filtration rate. Ann Intern Med 150, 604–612 (2009).

